# Trends and Determinants of Skilled Birth Attendants’ Use among Women of Reproductive Age in Tanzania: Evidence from the 2004/05-2022 National Surveys

**DOI:** 10.1101/2025.05.29.25328604

**Authors:** Jovin R. Tibenderana, Sanun Ally Kessy, Victoria Godfrey Majengo, Erick Donald Oguma, Tegemea Patrick Mwalingo, Mussa Hassan Bago, Immaculata P. Kessy, Elihuruma Eliufoo Stephano, Azan Abubakar Nyundo, Mtoro J. Mtoro

## Abstract

**Background:** Skilled birth attendants (SBAs) are vital for reducing maternal and newborn morbidity and mortality, yet their use remains below national and global targets, particularly in low—and middle-income countries like Tanzania. This study aimed to assess trends and determinants of SBA use among women of reproductive age (WRA) in Tanzania.

**Methods:** This was an analytical cross-sectional study among WRA in Tanzania, using data from the current Demographic and Health Surveys (DHS) 2004/05, 2010, 2015/16, and 2022. Data analysis was performed using STATA version 18. Analysis considered the complex survey design through the application of weights, clustering, and strata. Modified Poisson regression models estimated the determinants associated with SBA use among WRA. Results were presented using adjusted prevalence ratio (APR) with a 95% confidence interval.

**Results:** The study found an increased proportion of SBA use among WRA trends from 57.24% (2004/05) to 84.76% (2022), with the Dar es Salaam region showing higher SBA use of 86.6% in 2022. Residence (APR=0.96, 95% CI: 0.94, 0.99), education (APR=1.10, 95%CI: 1.04, 1.15), wealth index (APR= 1.07, 95% CI:1.02, 1.13), birth order (APR=0.91, 95% CI: 0.88, 0.93), distance to health facility (APR= 1.08, 95% CI: 1.05, 1.12), ANC visits (APR=1.13, 95% CI: 1.08, 1.17), and partners education (APR=1.15, 95% CI: 1.07, 1.23) were the determinants found significantly associated with SBA use.

**Conclusion:** Although SBA use has increased, national and global targets like 90% coverage by 2025 remain unmet. Efforts must focus on improving access, education, and male involvement, especially in rural areas. A comprehensive approach addressing both structural and cultural barriers is essential to ensure skilled care for all women during childbirth.

## Background

Having a Skilled Birth Attendant (SBA) present during childbirth positively contributes to reducing complications and improving birth outcomes, as they enable early identification and effective management of issues that may arise during delivery (1). SBA delivery refers to the assistance of childbirth by a trained healthcare provider, which is essential for ensuring the health and safety of both mothers and newborns, as the majority of maternal and newborn deaths occur during or immediately after birth(2). SBA delivery services serve as a crucial measure of access to high-quality maternal healthcare and is directly linked to Sustainable Development Goal (SDG) 3.1, which seeks to lower the global maternal mortality rate to fewer than 70 deaths per 100,000 live births by 2030 (3).

In 2023, an average of more than 700 women died each day from causes related to pregnancy and childbirth that could have been prevented (4). The World Health Organization (WHO) reported pregnancy-related complications in 2023, whereby around 260,000 women lost their lives during or after pregnancy and childbirth, and about 92% of these maternal deaths took place in low- and lower-middle-income countries (LMICs) that year (4,5). In Tanzania, the current Maternal Mortality Rate (MMR) stands at 104 per 100,000 live births (6)

Lack of access to SBA care during childbirth greatly raises the risk of complications for both mothers and newborns, such as excessive bleeding, infections, difficult labor, and lack of oxygen at birth (7,8). On a larger scale, low use of SBA highlights weaknesses in the healthcare system and hinders progress toward national and global health goals (9,10).

The Tanzanian government, working alongside development partners, has introduced several initiatives to improve access to SBA, which include expanding healthcare infrastructure, training and assigning skilled professionals, offering free maternal health services, and conducting community awareness programs (11). Policies such as the One Plan III and the Health Sector Strategic Plan V have focused on strengthening maternal health systems (12,13). Although these measures have led to some improvements, progress has been inconsistent, and a significant number of women, especially those in rural and remote areas, still give birth without the support of SBA. Tanzania’s national target was to achieve 90% coverage of births attended by skilled health workers by 2025 (12). Over time, progress has been made in the use of SBA. However, it has remained below national and global SBA coverage targets (2,12). Studies have shown that SBA use is associated with wealth index, residence (14–16), employment status (17), education (7,16,17), age(15), distance to health facility (7,18), media exposure(16,17), birth order (7,16), and ANC visit (7,16–18).

Despite various efforts, substantial gaps remain in understanding the determinants of SBA use in Tanzania. Existing studies are localized and lack rigorous, nationally representative analyses (7,10,19). Hence, to address this gap and to provide a comprehensive understanding of the SBA use, the present study aims to analyze trends and determinants of SBA use among women of reproductive age (WRA) in Tanzania using national representative data. The results of this study provide important information for health policymakers, program managers, and healthcare providers and guide efforts to improve maternal and neonatal health outcomes across the country.

## Methods and materials

### Study period and setting

The study used data from the 2004/05, 2010, 2015/16 and 2022 Demographic and Health Surveys (DHS), a comprehensive survey conducted every five years(6). Tanzania, East Africa’s largest nation, covers 940,000 square kilometers, including 60,000 square kilometers of inland water, and is situated south of the equator. It shares borders with eight countries: Kenya and Uganda to the north; Rwanda, Burundi, the Democratic Republic of Congo, and Zambia to the west; and Malawi and Mozambique to the south. As of 2022, Tanzania had an estimated population of 61,741,120 (20). The country is divided into thirty-one (31) Administrative regions, which are formally used by the Ministry of Health’s Reproductive and Child Health Section as a framework for organizing data.

### Study design and data source

This study was an analytical cross-sectional analysis utilizing nationally representative secondary data from the 2004/05, 2010, 2015/16 and 2022 Tanzania Demographic and Health Survey (TDHS), the publicly available datasets with extensive coverage of questions related to women’s data on skilled birth attendants (SBA). The TDHS is funded by the U.S. Agency for International Development (USAID), with implementation overseen by the Ministry of Health (MoH) in both Tanzania Mainland and Zanzibar, in collaboration with the National Bureau of Statistics (NBS) and the Office of the Chief Government Statistician (OCGS). Technical support for the initiative is provided by ICF International (20,21).

### Data collection procedure

The survey employs a standardized questionnaire, uniformly applied across all countries, to collect data from women aged 15–49. These questionnaires are translated into the main local languages of the participating nations. Before being finalized, the translated versions and the original English questionnaire undergo pretesting to ensure precision. During pretesting, field workers actively discuss the questions and suggest improvements to all versions. After the pretest, a debriefing session is conducted with the field staff, and the questionnaires are revised based on the feedback and insights gathered during this process (22).

### Sampling technique

The survey utilized multistage cluster sampling to collect data on various aspects of population health, such as neonatal mortality, health behaviors, nutritional status, family planning, and demographic characteristics. Initially, 629 clusters were identified, and households were selected. Within each cluster, 26 households were systematically chosen to ensure representation, leading to 16,354 households being included in the survey. Eligibility for participation was based on the presence of all women aged 15–49 years residing in the sampled household the night before the interview. Additional information about the sampling methodology and design has been documented in previous reports (6).

### Study variables

The outcome variable for this study examined whether a skilled birth attendant (SBA) was present during childbirth. In our analysis, doctors, nurses, midwives, and community SBAs were categorized as ‘skilled birth attendants’ and coded as ‘1’. Other attendants, including neighbors, relatives, traditional birth attendants (TBAs), and other workers, were classified as ‘unskilled birth attendants’ and coded as ‘0’. The coding aligned with previously published studies (17,23)

The independent variables were education, which was recategorized into no education, primary education, secondary education, or higher. Marital status was divided into married and unmarried. The wealth index was derived and regrouped into three equal subcategories: poor, middle, and rich. Employment status was classified into not working and working. Age categories were grouped into 15–24, 25–34, and 35–49 years. Distance to health care was categorized as a big problem and not a big problem. Media exposure was derived and classified into yes and no. Birth order was regrouped into 1 (first birth) and 2+ (the second or subsequent births). ANC visits were divided into <4 visits and ≥4 visits. Finally, partners’ education was recategorized into no education, primary education, secondary education, or higher education. The generation of variables, coding, categorization, and re-categorization were based on previous literature (14,17,24).

### Data management and analysis

Data management and statistical analyses were performed using STATA/MP 18.0 and Excel. Trends in the percentage of women who had SBA delivery and trends by administrative regions were plotted in Excel using data from the 2004/05, 2010, 2015/16, and 2022 TDHS. For normally distributed data, means with standard deviations (SD) were reported, while frequencies, percentages, and their confidence intervals were calculated for categorical variables to describe the study population’s characteristics. To address over- and under sampling issues, the sample was weighted (v005/1,000,000), and the survey set (svy) command in Stata was applied to account for the complex survey design.

Based on the Akaike Information Criterion (AIC) and Bayesian Information Criterion (BIC), modified Poisson regression models were used to identify determinants associated with SBA delivery using the 2022 DHS data. These models estimated the Prevalence Ratio (PR) and 95% confidence interval (CI) for enhanced precision of parameter estimates. The variables associated with crude regression were entered into adjusted analysis to identify independent variables associated with SBA delivery. Statistical significance was defined at a p-value of 0.05, and only variables significantly associated based on the Adjusted Prevalence Ratio (APR) were reported.

### Ethical clearance

Permission was granted to access and utilize the data via the DHS Program/ICF International website (http://www.dhsprogram.com). The data were only used for this study. The TDHS has released publications that provide complete information on the DHS program’s methodology and ethical criteria, which can be found elsewhere (6,25).

## RESULTS

### Social-demographic characteristics of the respondents in TDHS 2004/05-2022

Between 2004/05 and 2022, the socio-demographic profile of women of reproductive age in Tanzania showed notable shifts. The largest age group remained 25–34 years across all surveys, peaking at 45.78% in 2004/05. Rural residence consistently dominated, though urban representation rose from 22.1% in 2004/05 to nearly 30% in 2015/16 before slightly declining in 2022. Primary education was the most common level, but the proportion with secondary or higher education increased markedly from 5.2% (2004/05) to 23.3% (2022). Economic status remained relatively stable, with around 42% categorized as poor across years. While birth order trends shifted, women with two or more children surpassed first-time mothers only in 2022. Marriage rates declined from 74.5% (2004/05) to 58.1% (2022), alongside a drop in employment from 89.6% (2004/05) to 64.5% (2022). Perceived distance to health care as a big problem decreased notably in 2010, then rose again before falling to 33.4% in 2022. Media exposure fluctuated, with over half of the women lacking access in 2022. Antenatal care (≥4 visits) coverage increased over time, from 43.3% (2010) to 57.9% (2022).

**Table 1:** Percentage distribution of social-demographic characteristics of the respondents in TDHS 2004/05-2022 (N= 24,966, Weighted)

| Variable | Percentage n<br>(%) N=5772 | Percentage n<br>(%) N=5519 | Percentage n<br>(%) N=7079 | Percentage n<br>(%) N=6596 |
| --- | --- | --- | --- | --- |
| Years | 2004/05 | 2010 | 2015/16 | 2022 |
| **Age categories** |  |  |  |  |
| 15-24 | 1942 (33.63) | 1721 (31.18) | 2314 (32.69) | 2264 (34.32) |
| 25-34 | 2643 (45.78) | 2392 (43.35) | 2982 (42.13) | 2909 (44.11) |
| 35-49 | 1188 (20.59) | 1405 (25.47) | 1782 (25.18) | 1423 (21.57) |
| Mean (SD) | 28.92(7.23) | 29.65 (7.36) | 29.28(7.5) | 29 (7.0) |
| **Residence** |  |  |  |  |
| Urban | 1277 (22.12) | 1273 (23.07) | 2123 (29.99) | 1814 (27.5) |
| Rural | 4496 (77.88) | 4246 (76.93) | 4955 (70.01) | 4782 (72.5) |
| **Education** |  |  |  |  |
| No education | 1466 (25.4) | 1313 (23.79) | 1350 (19.08) | 1351 (20.49) |
| Primary | 4004 (69.36) | 3792 (68.72) | 4580 (64.7) | 3711 (56.26) |
| Secondary or higher | 302 (5.24) | 414(7.50) | 1149 (16.23) | 1534 (23.25) |
| **Wealth index** |  |  |  |  |
| Poor | 2413 (41.8) | 2309 (41.84) | 2947 (41.63) | 2797 (42.4) |
| Middle | 1166 (20.19) | 1175 (21.28) | 1349 (19.05) | 1285 (19.49) |
| Rich | 2194 (38.01) | 2035 (36.88) | 2783 (39.31) | 2514 (38.11) |
| **Birth order** |  |  |  |  |
| 1 | 3246 (56.23) | 3213 (58.22) | 4472 (63.17) | 3278 (49.7) |
| 2+ | 2527 (43.75) | 2306 (41.78) | 2607 (36.83) | 3318 (50.3) |
| **Marital status** |  |  |  |  |
| Unmarried | 1470 (25.46) | 1298 (23.53) | 3007 (42.48) | 2764 (41.91) |
| Married | 4303 (74.54) | 4220 (76.47) | 4071 (57.52) | 3832 (58.09) |
| **Employment status** |  |  |  |  |
| Not working | 599 (10.37) | 652 (11.81) | 1142 (16.13) | 2339 (35.46) |
| Working | 5174 (89.63) | 4867 (88.19) | 5937 (83.87) | 4257 (64.54) |
| **Distance to health care** |  |  |  |  |
| Big problem | 2309 (40) | 1213 (21.97) | 3226 (45.58) | 2202 (33.38) |
| Not a big problem | 3463 (60) | 4306 (78.03) | 3852 (54.42) | 4394 (66.62) |
| **Media exposure** |  |  |  |  |
| No | 2408 (41.72) | 2124 (38.5) | 3368 (47.59) | 3447 (52.26) |
| Yes | 3364 (58.28) | 3394 (61.5) | 3710 (52.41) | 3149 (47.74) |
| **ANC visits** |  |  |  |  |
| <4 | 2202(38.16) | 3129(56.69) | 3456(48.81) | 2774 (42.06) |
| ≥4 | 3570 (61.84) | 2390 (43.31) | 3623 (51.19) | 3822 (57.94) |
| **Partner's Education** |  |  |  |  |
| No education | 1470 (24.47) | 1211 (21.94) | 1234 (17.43) | 1241 (18.81) |
| Primary | 3508 (60.78) | 3453 (62.57) | 4321 (61.04) | 3587 (54.38) |
| Secondary or higher | 794 (14.75) | 855(15.49) | 1524 (21.53) | 1768 (26.81) |

### Trends of SBA use from 2004/05 to 2022

The use of SBA among women of reproductive age in Tanzania showed a fluctuating but overall upward trend between 2004/05 and 2022. In 2004/05, SBA use was 57.24%, slightly declining to 54.43% in 2010. However, this was followed by a significant rise to 67.28% in 2015 and further to 84.76% in 2022 (**Figure 1**)

**Figure 1:**
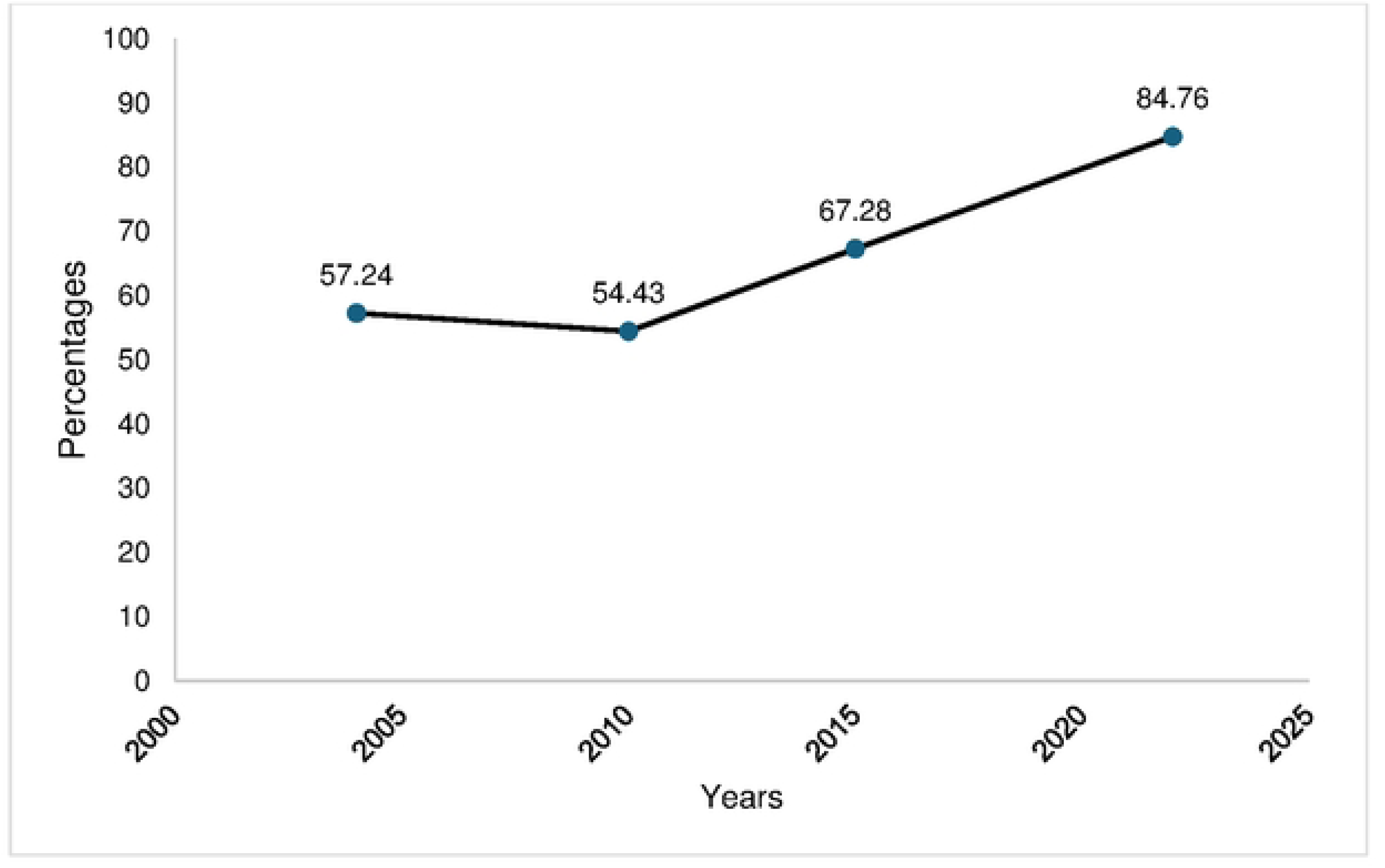
Trends of SBA use among WRA 2004/05-2022

### Trend of SBA use by regions from 2004/05 to 2022

Between 2004/05 and 2022, the use of SBA among women of reproductive age showed varied trends across Tanzanian regions. Dar es Salaam consistently recorded the highest SBA use, peaking at 95.5% in 2015 before declining to 86.6% in 2022, still maintaining a high utilization level. Kilimanjaro and Iringa also maintained high SBA coverage above 80% from 2004/05 to 2022. Regions like Dodoma and Arusha exhibited substantial improvements, with Dodoma rising from 61.36% in 2004/05 to 83.6% in 2022, and Arusha increasing from 60.98% to 76.85% over the same period. Tabora and Simiyu showed remarkable progress, with SBA use jumping from 56.6% and 40.79% in 2004/05 to 78.87% and 78.53% in 2022, respectively (**Figure 2**)

**Figure 2:**
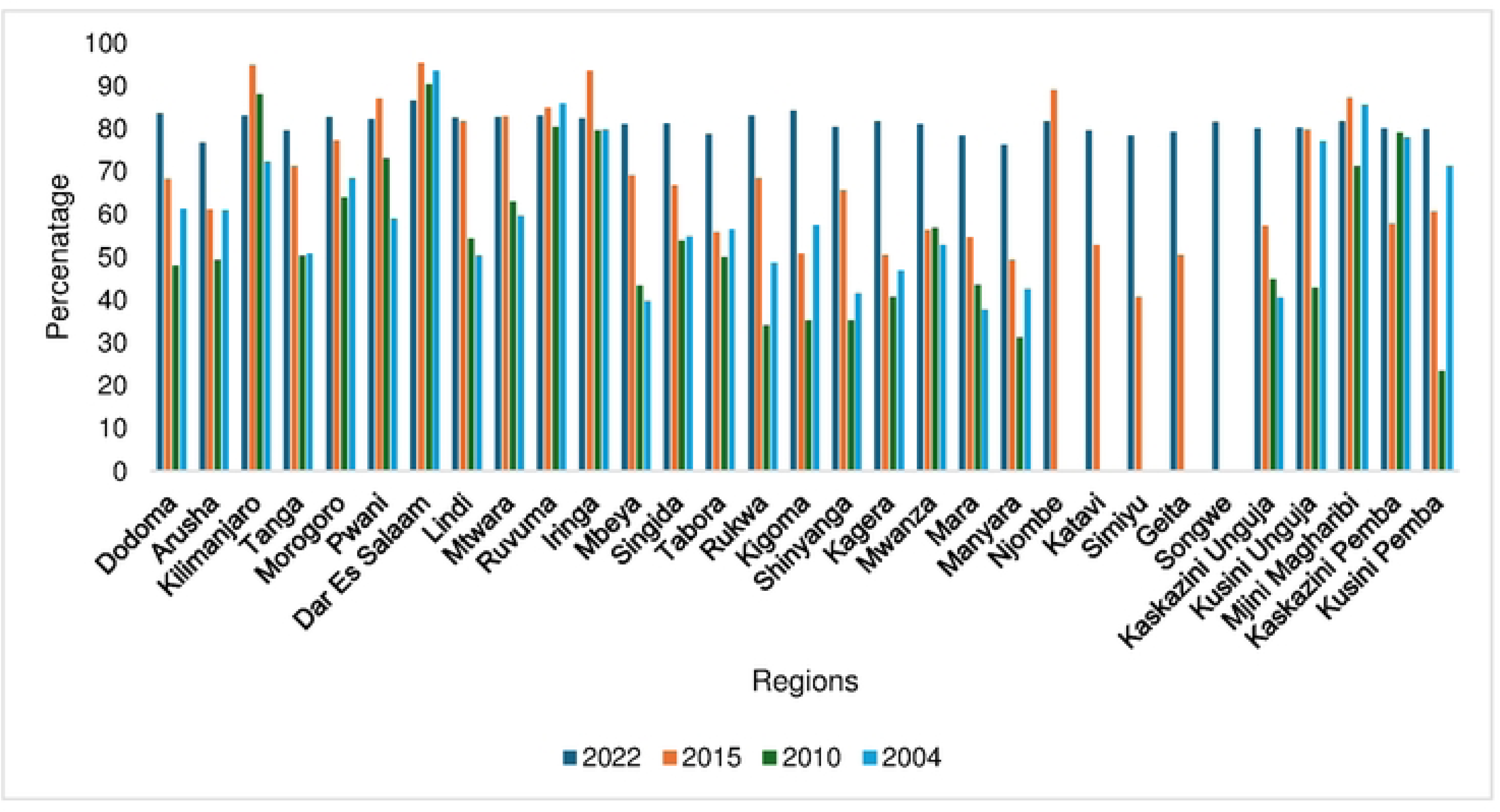
Trends of SBA use among WRA by regions, 2004/05-2022

### Factors associated with SBA use during childbirth among WRA

After adjusting for other factors, WRA who were residing in rural areas had less prevalence of SBA use compared to their counterparts (APR=0.96, 95% CI: 0.94, 0.99), higher prevalence of SBA use was observed among WRA who had secondary or higher education (APR=1.10, 95%CI: 1.04, 1.15). Women who had more than 4 ANC visits exhibited a 13% higher prevalence of SBA use than those who didn’t (APR=1.13, 95% CI: 1.08, 1.17). Lastly, women whose partner had secondary or higher education exhibited a higher prevalence of SBA use compared to those who didn’t have formal education (APR=1.15, 95% CI: 1.07, 1.23)

**Table 2:** Factors associated with SBA use during childbirth among WRA.

| Variable | CPR (95% CI) | P-value | APR (95% CI) | P-value |
| --- | --- | --- | --- | --- |
| **Age categories** |  |  |  |  |
| 15-24 | 1 |  | 1 |  |
| 25-34 | 0.99(0.96, 1.02) | 0.588 | 0.99(0.97, 1.03) | 0.898 |
| 35-49 | 0.96(0.93, .99) | 0.025 | 1.00(0.96, 1.04) | 0.908 |
| **Residence** |  |  |  |  |
| Urban | 1 |  | 1 |  |
| Rural | 0.84(0.83, 0.86) | <0.001 | 0.96(0.94, 0.99) | 0.017 |
| **Education** |  |  |  |  |
| No education | 1 |  | 1 |  |
| Primary | 1.18(1.13, 1.24) | <0.001 | 1.07(1.02, 1.12) | 0.004 |
| Secondary or higher | 1.32(1.27, 1.38) | <0.001 | 1.10(1.04, 1.15) | <0.001 |
| **Wealth index** |  |  |  |  |
| Poor | 1 |  | 1 |  |
| Middle | 1.16(1.12, 1.20) | <0.001 | 1.08(1.03, 1.12) | <0.001 |
| Rich | 1.27(1.24, 1.31) | <0.001 | 1.07(1.02, 1.13) | 0.003 |
| **Birth order** |  |  |  |  |
| 1 | 1 |  |  |  |
| 2+ | 0.86(0.84, 0.89) | <0.001 | 0.91(0.88, 0.93) | <0.001 |
| **Marital status** |  |  |  |  |
| Unmarried | 1 |  |  |  |
| Married | 0.97(0.95, 1.00) | 0.055 |  |  |
| **Employment status** |  |  |  |  |
| Not working | 1 |  |  |  |
| Working | 1.02(0.98, 1.05) | 0.249 |  |  |
| **Distance to health care** |  |  |  |  |
| Big problem | 1 |  | 1 |  |
| Not a big problem | 1.20(1.16, 1.24) | <0.001 | 1.08(1.05, 1.12) | <0.001 |
| **Media exposure** |  |  |  |  |
| No | 1 |  | 1 |  |
| Yes | 1.13(1.09, 1.16) | <0.001 | 1.01(0.98, 1.04) | 0.483 |
| **ANC visits** |  |  |  |  |
| <4 | 1 |  | 1 |  |
| ≥4 | 1.21(1.16, 1.26) | <0.001 | 1.13(1.08, 1.17) | <0.001 |
| **Partner's Education** |  |  |  |  |
| No education | 1 |  | 1 |  |
| Primary | 1.26(1.18, 1.34) | <0.001 | 1.12(1.05, 1.20) | 0.001 |
| Secondary or higher | 1.40(1.32, 1.49) | <0.001 | 1.15(1.07, 1.23) | <0.001 |

## DISCUSSION

The study aimed to determine trends and determinants associated with SBA use among WRA in Tanzania from 2004/05 to 2022. The study found an overall increase in the proportion of SBA use among WRA trends from 57.24% to 84.76% from 2004/05 to 2022, respectively, with urbanized regions like Dar es Salaam showing higher SBA use. Residence, education, wealth index, birth order, distance to health facility, ANC visits, and partners’ education were the determinants found significantly associated with SBA The findings reveal a significant upward trend in the use of SBA among WRA in Tanzania between 2004/05 and 2022. This gradual rise indicates significant advancements in access to and delivery of maternal healthcare, which can likely be attributed to interventions by the government and its partners. These efforts include providing free maternal care, expanding health facilities, and conducting community awareness campaigns (11). However, despite this positive progress, achieving the national goal of 90% SBA coverage by 2025 (12) (which is the current year) may still pose challenges. Ongoing barriers to access, particularly in rural and underserved regions, as witnessed from findings from this study, along with inequalities related to socioeconomic status and education, could limit the realization of SBA use.

As previously reported, (8,14) the negative association between rural residence and SBA use reflects persistent disparities in maternal healthcare access. Women in rural areas face challenges such as limited availability of health facilities, poor transportation infrastructure, and lower exposure to health education (26,27). Additionally, cultural preferences for traditional birth attendants and higher levels of poverty and illiteracy contribute to the underutilization of skilled services (28,29). These barriers hinder rural women from accessing timely and adequate care during childbirth.

Being educated increased the chances of SBA use. This can be explained by the fact that education plays a key role in improving women’s health knowledge, helping them recognize the value of professional care during childbirth and enabling them to make informed choices (30). Women with higher levels of education are also more likely to have economic independence, which increases their access to healthcare services and their ability to decide when and where to seek care (31). Additionally, advanced education is often linked to later marriage and childbearing, improving maternal health outcomes. This study aligns with previously published studies (7,16,17).

Women who attended ≥ 4 ANC visits had a higher prevalence of SBA use. Regular ANC visits provide women with more opportunities to receive health education, prepare for childbirth, and build confidence in the healthcare system (32). During these visits, healthcare providers also offer counseling on the advantages of delivering with a skilled attendant and may refer expectant mothers to suitable facilities(33). This strong connection highlights the significance of encouraging consistent ANC attendance as a vital approach to increasing the use of skilled birth attendants and improving the health outcomes of both mothers and newborns. Similar findings were reported by previous studies (7,16–18).

Findings from this study have shown that a partner’s level of education plays a key role in determining whether women of reproductive age use skilled birth attendants (SBA), i.e women whose partners have completed secondary school or higher are more likely to give birth with the help of an SBA compared to those whose partners have less education. The reasons behind this could be that better-educated partners tend to have greater knowledge about maternal health, are more supportive of seeking medical care, and are more likely to encourage deliveries at health facilities (34). These findings align with the study done in India (35).

As supported by previous evidence (14–16), women from rich households were found to be more likely to use skilled birth attendants (SBA) during delivery compared to those in the lowest wealth group. This can be explained by several factors, including limited support for accessing SBA services among poorer women, who may have greater household or family responsibilities. Financial barriers also play a role, as low-income households may struggle with transportation costs to reach health facilities or afford essential supplies needed during childbirth. Although SBA delivery services are provided free at public health facilities, mothers are sometimes required to purchase necessary items that may not be available at the health facility at the time of delivery (36,37).

### Strengths and Limitations

A key strength of this study is the use of the Demographic and Health Survey (DHS) data, which is widely recognized for its rigorous methodology and has been validated in numerous prior studies. The nationally representative nature of the dataset enhances the generalizability of our findings, not only to WRA in Tanzania but also to those in other African countries with similar contexts. However, the study has several limitations. As it relies on secondary data, our analysis was limited to the variables included in the dataset; that is, including essential factors such as cultural beliefs or patriarchal norms that may influence women’s use of SBA was impossible. Furthermore, the cross-sectional design of the DHS does not allow for causal inferences to be drawn. Additionally, while the large sample size boosts statistical power, it may lead to even minor associations being statistically significant, which could affect the interpretation of effect sizes. Despite these limitations, our findings offer meaningful insights that can inform future research and support the development of targeted public health interventions.

### Conclusion and recommendation

Although using SBA among WRA has shown a positive upward trend, national and global targets such as the 90% target by this year (2025) remain unmet. These findings underscore the urgent need for continued and targeted efforts to increase SBA utilization to advance progress toward Sustainable Development Goal 3.1. To achieve this, it is essential to implement targeted interventions that improve access to and awareness of SBA services, particularly in rural areas where utilization remains low. Strengthening women’s education is also critical, as it significantly influences the uptake of maternal healthcare services. Additionally, engaging men in maternal health decisions and fostering joint decision-making within couples can play a transformative role in improving SBA use. A multifaceted approach that addresses both structural and socio-cultural barriers is vital to ensure that every woman has access to skilled care during childbirth.

## Data Availability

The data underlying the results presented in the study are available from http://www.dhsprogram.com

## Acknowledgments

The authors are grateful to MEASURE DHS for providing them with the data set.

## Abbreviations

AIC: Akaike Information
BIC: Bayesian Information Criteria
CI: Confidence Interval
DHS: Demographic and Health Survey
EA: Enumeration Area
LMICs: low- and lower-middle-income countries
MMR: Maternal Mortality Rate
NBS: National Bureau of Statistics
PR: Prevalence Ratio
PSU: Primary Sampling Unit
WRA: Women of Reproductive Age
SBA: Skilled Birth Attendants
TDHS: Tanzania Demographic and Health Survey

